# APOE and amyloid-tau pathology in cognitively unimpaired older adults

**DOI:** 10.64898/2026.06.02.26354753

**Authors:** Carlos Albarrán Morillo, Lukai Zheng, Elham Ghanbarian, Babak Khorsand, Crystal M. Glover, Joshua D. Grill, S. Ahmad Sajjadi, Alzheimer’s Disease Neuroimaging Initiative, Ali Ezzati

## Abstract

**INTRODUCTION:** APOE genotype shows well-established dose-dependent associations with higher amyloid in cognitively unimpaired (CU) adults. In contrast, associations with tau burden and cognition are less well characterized.

**METHODS:** We performed a cross-sectional analysis of harmonized multi-cohort ADSP-PHC data from 4,380 CU participants across 4 cohorts with APOE genotype, amyloid PET, and cognitive data from four domains of memory, language, executive, and visuospatial function, including a subset of 758 with tau PET imaging.

**RESULTS:** APOE ε4 showed a strong dose-dependent association with amyloid burden and amyloid positivity, with the highest levels observed among ε4 homozygotes. Associations between APOE and global tau burden were more modest and appeared to be driven mainly by ε4 homozygotes, while regional analyses showed localized APOE ε4-related associations in medial temporal regions. Independently, higher tau burden was associated with lower memory and language performance.

**CONCLUSIONS:** In CU older adults, APOE ε4 was most strongly associated with amyloid burden, with more modest associations observed for medial temporal tau burden.

**Highlights:**

- APOE ε4 showed dose-dependent associations with amyloid burden and age-related amyloid differences
- APOE-related global tau associations were modest, mainly in ε44 group, with stronger medial temporal effects among ε4 carriers
- Higher tau burden was associated with lower memory and language performance, regardless APOE status

**RESEARCH IN CONTEXT:**

1. **Systematic review:** The authors reviewed the literature using PubMed and Google Scholar to examine relationships among APOE genotype, amyloid pathology, tau pathology, and cognition in cognitively unimpaired individuals. Previous studies from A4, ADNI, A4/LEARN, and large multicohort tau PET datasets have consistently shown strong APOE-related effects on amyloid burden and more modest regional effects on tau. The present study extends this literature using a large harmonized ADSP-PHC dataset with amyloid PET, tau PET, and harmonized cognitive measures.
2. **Interpretation:** The findings suggest that amyloid abnormalities remain the main pathological feature linked to APOE genotype in cognitively unimpaired individuals. APOE-related effects on tau were smaller and were concentrated mainly in medial temporal regions. APOE genotype was not consistently related to cognition, while higher tau burden was associated with lower memory and language performance.
3. **Future directions:** These findings support the use of APOE genotype as a stratification factor in studies of cognitively unimpaired individuals. Longitudinal studies will be important to clarify how APOE-related differences in amyloid and regional tau burden evolve over time and whether they are linked to later cognitive decline and progression to Alzheimer’s disease.

## Introduction

APOE ε4 is the strongest genetic risk factor for sporadic Alzheimer’s disease (AD) [1,2]. Carrying one APOE ε4 allele is associated with an approximately 2-to 3-fold higher risk of AD, while individuals with two ε4 alleles may have a 10–15-fold greater risk and develop dementia at an earlier age, supporting a dose-dependent effect [3]. In contrast, the APOE ε2 allele is generally associated with lower AD risk and later onset of AD-related biomarker changes. ε2 carriers have an estimated odds ratio of about 0.6, compared to individuals with ε33, the most common APOE genotype [4]. APOE and its protein product, apoE, are involved in several biological pathways linked to AD, including amyloid-β (Aβ) aggregation, fibril formation, and clearance [5]. Consistent with these effects, APOE ε4 carriers typically become amyloid positive about 10–15 years earlier than ε33 carriers, in contrast amyloid positivity tends to occur later among ε2 carriers [6].

Recent studies suggest that APOE may also contribute to tau accumulation and downstream neurodegeneration. Because Aβ and tau pathology are closely linked in AD [7], part of the association between APOE and tau may be explained by the effect of APOE on amyloid accumulation. However, whether APOE is also associated with tau pathology and subtle domain-specific cognitive changes independent of amyloid burden remains uncertain, particularly in cognitively unimpaired (CU) individuals. Findings across studies have remained inconsistent. Lowe and colleagues [8] reported that medial temporal tau PET abnormalities in CU individuals were associated primarily with age and amyloid positivity, with no significant association observed for APOE status. Ramanan and colleagues [9] similarly found limited APOE ε4-related tau effects after adjustment for global amyloid levels. In contrast, Ossenkoppele and colleagues [10] identified APOE ε4 carrier status as a factor associated with tau PET positivity, and Ghisays and colleagues [11] showed that APOE ε4 modified the relationship between age and entorhinal tau deposition. Young and colleagues [12] further reported that among amyloid-positive CU individuals, APOE ε4 was associated with higher regional tau PET burden even after accounting for cross-sectional amyloid burden. In a separate multicohort study, Ossenkoppele and colleagues [13] showed that tau PET positivity in temporal composite regions increased with APOE ε4 carrier status in CU individuals in models accounting for amyloid status. Weigand and colleagues [14] also found that APOE ε4 interacted with medial temporal lobe (MTL) tau pathology to influence memory performance independently of amyloid burden in older adults without dementia. Across these studies, APOE-related tau effects tended to be more evident in analyses restricted to amyloid-positive individuals because in mixed amyloid-negative and amyloid-positive samples, stronger amyloid-related effects may attenuate smaller APOE-related tau associations.

Given these mixed findings, we used harmonized data from 4 cohorts within the Alzheimer’s Disease Sequencing Project Phenotype Harmonization Consortium (ADSP-PHC) to examine associations among APOE genotype, amyloid burden, tau pathology, and cognition in CU older adults. We performed a cross-sectional analysis of 4,380 individuals with amyloid PET data, including a subset of 758 individuals with tau PET data. First, we evaluated dose-related associations between APOE genotype and amyloid burden. Second, we examined the relative contributions of amyloid burden and APOE genotype to global and regional tau pathology, including analyses restricted to amyloid-positive individuals. Third, we assessed associations between tau burden and domain-specific cognitive performance. We hypothesized that APOE ε4 would show the strongest association with amyloid burden, that amyloid burden would be the primary correlate of tau pathology with smaller APOE-related tau effects, and that higher tau burden would be associated with lower domain-specific cognitive performance, particularly memory, independently of APOE status.

## Methods

### Study design and participants

This cross-sectional study used data from ADSP-PHC Release 3 [15], which includes harmonized demographic, cognitive, imaging, and biomarker data collected across multiple cohorts. The ADSP-PHC was established to standardize and harmonize phenotypic data across studies, enabling large-scale pooled analyses and supporting integrative investigations of genetic, imaging, biomarker, and cognitive factors related to Alzheimer’s disease.

Participant selection and study inclusion are summarized in Figure S1. Among 64,932 participants available in ADSP-PHC Release 3 extraction used for this study, 7,128 were excluded because APOE genotype data were unavailable. Of these, 51,330 did not have amyloid PET data, resulting in 6,474 individuals with available amyloid imaging. The analytic sample was subsequently restricted to CU participants, excluding 1,348 individuals with cognitive impairment, resulting in 5,126 participants. Criteria used to define CU status in the contributing cohorts are briefly described in the following section. An additional 714 participants were excluded because cognitive data were unavailable across all domains, and 32 were excluded because the interval between amyloid PET and cognitive assessment exceeded 1 year. The final amyloid PET sample included 4,380 participants. Most participants came from the A4 Study (n = 3,310), Alzheimer’s Disease Neuroimaging Initiative (ADNI; n = 691), Wisconsin Registry for Alzheimer’s Prevention (WRAP; n = 236), and NIA Alzheimer Disease Centers (ADC; n = 143).

For the tau PET subset, 3,586 participants without tau PET (^18^F-flortaucipir [FTP]) data were excluded, leaving 794 individuals. Another 36 participants were excluded because the interval between amyloid PET and tau PET exceeded 1 year, resulting in a final tau PET subset of 758 participants. This subset consisted primarily of participants from ADNI (n = 421), the A4 Study (n = 333), and ADC (n = 4).

All contributing studies received approval from their local institutional review boards, and all participants provided written informed consent.

### Clinical and cognitive assessments

Clinical and cognitive data were obtained from ADSP-PHC datasets. Neuropsychological measures collected across participating cohorts were harmonized using previously established ADSP-PHC procedures [16]. Briefly, individual test items were mapped to cognitive domains, co-calibrated across studies using common anchor measures, and combined using factor-analytic models to generate harmonized scores for memory, language, executive functioning, and visuospatial functioning [16]. These scores are standardized, with higher values indicating better cognitive performance. Because neuropsychological batteries differed across cohorts, the availability of domain scores varied by study. Notably, the A4 Study contributed harmonized memory scores but did not contribute scores for the other cognitive domains; therefore, analyses of language, executive functioning, and visuospatial functioning were based on smaller subsets and may be more influenced by cohort composition and missing data patterns. Analyses focused on domain-specific cognitive measures rather than a global cognitive composite.

Cognitive status classifications were assigned by the individual cohorts and were not harmonized by ADSP-PHC. In the A4 Study, participants were required to be cognitively unimpaired at enrollment, with a Clinical Dementia Rating (CDR) score of 0 and normal cognitive performance despite elevated amyloid burden. In ADNI, cognitively unimpaired participants were defined by a CDR score of 0, normal memory performance, preserved daily functioning, and the absence of dementia. WRAP classified participants using longitudinal neuropsychological and clinical evaluations, with cognitively unimpaired individuals showing no evidence of mild cognitive impairment or dementia. NIA Alzheimer Disease Centers (ADC) assigned cognitive status using standardized clinical and neuropsychological assessments, classifying cognitively unimpaired participants as those who did not meet criteria for mild cognitive impairment or dementia. Only participants classified as cognitively unimpaired (PHC_Diagnosis = 1) were included in the present analyses.

### Amyloid and tau PET imaging

Amyloid and tau PET measures were obtained from the ADSP-PHC MRI-free PET processing pipeline, which provides standardized PET quantification across cohorts through template-based image normalization and regional uptake extraction [17]. Only scans that passed quality control procedures and met predefined post-injection acquisition windows were included to improve consistency across tracers and cohorts. Amyloid burden was quantified using standardized uptake value ratios (SUVRs) derived from a global cortical composite and normalized to the whole cerebellum reference region. SUVRs were converted to Centiloid (CL) units using tracer-specific transformations to facilitate comparisons across amyloid PET ligands [18]. Amyloid positivity was defined using a global threshold of CL ≥ 25, consistent with previously reported Centiloid thresholds for elevated amyloid burden [19].

Tau pathology was measured using SUVRs normalized to an inferior cerebellar gray matter reference region. Global tau burden was represented by a meta-temporal composite calculated as a volume-weighted average across bilateral entorhinal, amygdala, parahippocampal, fusiform, inferior temporal, and middle temporal regions, consistent with Braak stage–related patterns of tau deposition [20]. Tau PET imaging was performed using 18F-flortaucipir (FTP).

Additional regional analyses were performed to evaluate whether associations varied across individual regions contributing to the meta-temporal composite and selected neocortical regions. These included the entorhinal cortex, amygdala, inferior temporal cortex, inferior parietal cortex, and precuneus [12].

### Data analysis

Regression models were used to examine associations among APOE genotype, amyloid burden, tau pathology, and cognitive performance. Continuous outcomes, including Centiloid values, global and regional tau burden, and domain-specific cognitive scores, were analyzed using linear regression. Amyloid positivity was analyzed using logistic regression.

Initial analyses focused on the associations between APOE genotype and amyloid burden using both continuous Centiloid measures and binary amyloid positivity. We then examined associations between amyloid burden and global tau pathology using both amyloid positivity and continuous Centiloid values. Associations between APOE and global tau burden were subsequently tested after adjusting for continuous amyloid burden. The same analyses were repeated in amyloid-positive individuals only. APOE effects were evaluated using APOE genotype, ε4 carrier status, and ε4 homozygous status. These analyses were then extended to regional tau measures. Multiple comparisons across regional tau analyses were controlled using false discovery rate (FDR) correction, and both uncorrected and FDR-corrected p values are reported.

APOE × age and APOE × sex interactions were tested to evaluate age- and sex-related differences in amyloid and tau burden. Additional sensitivity analyses used spline-based age models to examine possible nonlinear age effects, including APOE ε4 × spline(age) interaction terms. Model fit was compared using likelihood ratio tests (LRTs) and changes in Akaike information criterion (ΔAIC).

Relationships between tau burden and domain-specific cognitive performance were examined using all available cognitive domain data. Sensitivity analyses included robust and quantile regression models to evaluate the consistency of findings across different model specifications and across the distribution of cognitive performance.

All models were adjusted for age, sex, education, and study cohort. Initial analyses modeled APOE genotype as a six-category categorical variable (ε22, ε23, ε33, ε24, ε34, and ε44), with ε33 as the reference group. When overall genotype associations were not significant, additional analyses evaluated APOE ε4 carrier status and APOE ε44 homozygous status separately. Age and sex interaction terms were retained only when statistically supported; otherwise, reduced additive models were used.

Group differences were assessed using Kruskal–Wallis tests with Dunn post hoc comparisons and Holm correction. Categorical variables were analyzed using chi-square tests. Statistical significance was defined as a two-sided P < .05.

All analyses were performed in Python using standard scientific libraries.

## Results

### Sample Characteristics

Baseline characteristics are summarized in Tables 1 and 2. The amyloid PET sample included 4,380 participants with a mean (SD) age of 71.3 (5.6) years; 60.1% were women, and the mean (SD) years of education was 16.7 (2.8). The most common APOE genotype was ε33 (54.6%), followed by ε34 (29.3%) and ε23 (10.2%). Smaller proportions included ε24 (2.4%), ε44 (3.2%), and ε22 (0.3%).

**Table 1.** Participant characteristics of the amyloid PET analytic sample, overall and by APOE genotype.

| Characteristic | Overall | ε22 | ε23 | ε33 | ε24 | ε34 | ε44 | p-value |
| --- | --- | --- | --- | --- | --- | --- | --- | --- |
| N (%) | 4,380 | 14 (0.3) | 447 (10.2) | 2,390 (54.6) | 107 (2.4) | 1,282 (29.3) | 140 (3.2) |  |
| Age, mean (SD), y | 71.3 (5.6) | 69.63 (4.12) | 72.06 (5.71) | 71.66 (5.74) | 71.53 (5.65) | 70.48 (5.36) | 69.92 (5.36) | < 0.001 |
| Women, N (%) | 2,631 (60.1) | 8 (57.1) | 257 (57.5) | 1,450 (60.7) | 66 (61.7) | 771 (60.1) | 79 (56.4) | 0.771 |
| Race, N (%) |  |  |  |  |  |  |  |  |
| American Indian or Alaska Native | 13 (0.3) | 0 | 0 | 6 (0.3) | 0 | 6 (0.5) | 1 (0.7) |  |
| Asian | 73 (1.7) | 0 | 4 (0.9) | 57 (2.4) | 0 | 11 (0.9) | 1 (0.7) |  |
| Black or African American | 203 (4.6) | 0 | 43 (9.6) | 89 (3.7) | 8 (7.5) | 54 (4.2) | 9 (6.4) |  |
| Native Hawaiian or Other Pacific Islander | 2 (0.0) | 0 | 0 | 1 (0.0) | 0 | 1 (0.1) | 0 |  |
| White | 4,028 (92.0) | 14 (100.0) | 395 (88.4) | 2,199 (92.0) | 97 (90.7) | 1,195 (93.2) | 128 (91.4) |  |
| Other/Unknown/Multiple | 61 (1.4) | 0 | 5 (1.1) | 38 (1.6) | 2 (1.9) | 15 (1.2) | 1 (0.7) |  |
| Ethnicity, N (%) |  |  |  |  |  |  |  |  |
| Hispanic or Latino | 176 (4.0) | 0 | 9 (2.0) | 115 (4.8) | 5 (4.7) | 42 (3.3) | 5 (3.6) | 0.048 |
| Not Hispanic or Latino | 4,180 (96.0) | 14 (100.0) | 435 (98.0) | 2262 (95.2) | 101 (95.3) | 1,233 (96.7) | 135 (96.4) |  |
| Education, mean (SD), y | 16.7 (2.8) | 16.86 (2.14) | 16.60 (3.01) | 16.60 (2.72) | 16.60 (2.99) | 16.80 (2.69) | 16.77 (2.55) | 0.699 |
| Memory, mean (SD) | 0.74 (0.44) | 0.64 (0.32) | 0.73 (0.42) | 0.75 (0.45) | 0.71 (0.44) | 0.72 (0.43) | 0.72 (0.46) | 0.502 |
| Executive Function, mean (SD) | 0.86 (0.60) | – | 0.89 (0.56) | 0.87 (0.60) | 0.74 (0.53) | 0.86 (0.63) | 0.73 (0.50) | 0.562 |
| Language, mean (SD) | 0.85 (0.56) | – | 0.79 (0.49) | 0.85 (0.57) | 0.82 (0.48) | 0.87 (0.57) | 0.87 (0.50) | 0.822 |
| Visuospatial, mean (SD) | 0.49 (0.49) | – | 0.49 (0.46) | 0.48 (0.51) | 0.43 (0.51) | 0.52 (0.47) | 0.57 (0.39) | 0.804 |
| Aβ positive, N (%) | 1,300 (29.7) | 2 (14.3) | 76 (17.0) | 454 (19.0) | 52 (48.6) | 614 (47.9) | 102 (72.9) | < 0.001 |
| Aβ SUVR (Centiloids), mean (95% CI) | 17.91 (16.89–18.93) | 4.64 (–2.92–12.20) | 6.78 (4.46–9.09) | 9.73 (8.59–10.87) | 28.15 (21.70–34.60) | 32.47 (30.32–34.63) | 53.20 (46.40–60.00) | < 0.001 |

Abbreviations: APOE, apolipoprotein E; SD, standard deviation; CI, confidence interval; Aβ, amyloid-β; SUVR, standardized uptake value ratio; –, missing data.

**Table 2.** Participant characteristics of the tau PET analytic sample, overall and by APOE genotype.

| Characteristic | Overall | ε22 | ε23 | ε33 | ε24 | ε34 | ε44 | p-value |
| --- | --- | --- | --- | --- | --- | --- | --- | --- |
| N (%) | 758 | 1 (0.1) | 62 (8.2) | 370 (48.8) | 20 (2.6) | 277 (36.5) | 28 (3.7) |  |
| Age, mean (SD), y | 72.3 (6.1) | 68.73 | 72.65 (6.03) | 73.18 (6.39) | 72.84 (5.58) | 71.09 (5.42) | 70.89 (5.82) | 0.002 |
| Women, N (%) | 456 (60.2) | 1 (100.0) | 36 (58.1) | 214 (57.8) | 11 (55.0) | 173 (62.5) | 21 (75.0) | 0.414 |
| Race, N (%) |  |  |  |  |  |  |  |  |
| American Indian or Alaska Native | 3 (0.4) | 0 | 0 | 2 (0.5) | 0 | 1 (0.4) | 0 |  |
| Asian | 14 (1.8) | 0 | 1 (1.6) | 11 (3.0) | 0 | 2 (0.7) | 0 |  |
| Black or African American | 36 (4.7) | 0 | 8 (12.9) | 13 (3.5) | 1 (5.0) | 14 (5.1) | 0 |  |
| White | 691 (91.2) | 1 (100.0) | 52 (83.9) | 335 (90.5) | 18 (90.0) | 258 (93.1) | 27 (96.4) |  |
| Other/Unknown/Multiple | 14 (1.8) | 0 | 1 (1.6) | 9 (2.4) | 1 (5.0) | 2 (0.7) | 1 (3.6) |  |
| Ethnicity, N (%) |  |  |  |  |  |  |  |  |
| Hispanic or Latino | 28 (3.7) | 0 | 2 (3.2) | 15 (4.1) | 0 | 11 (4.0) | 0 | 0.830 |
| Not Hispanic or Latino | 724 (96.3) | 1 (100.0) | 60 (96.8) | 353 (95.9) | 20 (100.0) | 262 (96.0) | 28 (100.0) |  |
| Education, mean (SD), y | 16.6 (2.5) | 18.00 | 16.37 (2.54) | 16.82 (2.50) | 15.80 (3.00) | 16.34 (2.44) | 16.64 (2.86) | 0.194 |
| Memory, mean (SD) | 0.81 (0.49) | 0.89 | 0.80 (0.47) | 0.86 (0.49) | 0.63 (0.44) | 0.76 (0.48) | 0.79 (0.55) | 0.055 |
| Executive Function, mean (SD) | 0.91 (0.55) | – | 0.86 (0.47) | 0.94 (0.56) | 0.47 (0.49) | 0.94 (0.55) | 0.68 (0.60) | 0.140 |
| Language, mean (SD) | 0.86 (0.55) | – | 0.75 (0.50) | 0.84 (0.54) | 0.75 (0.31) | 0.95 (0.61) | 0.75 (0.37) | 0.194 |
| Visuospatial, mean (SD) | 0.47 (0.51) | – | 0.48 (0.44) | 0.45 (0.52) | 0.16 (0.64) | 0.53 (0.51) | 0.33 (0.44) | 0.273 |
| Aβ positive, N (%) | 384 (50.7) | 1 (100.0) | 17 (27.4) | 139 (37.6) | 14 (70.0) | 190 (68.6) | 23 (82.1) | < 0.001 |
| Aβ SUVR (Centiloids), mean (95% CI) | 32.95 (30.14–35.76) | 28.00 | 10.29 (4.44–16.14) | 22.46 (18.84–26.08) | 36.40 (23.70–49.10) | 49.01 (44.28–53.73) | 60.61 (44.18–77.04) | < 0.001 |
| Tau SUVR, mean (95% CI) | 1.24 (1.23–1.25) | 1.18 | 1.21 (1.19–1.23) | 1.23 (1.22–1.25) | 1.23 (1.17–1.28) | 1.26 (1.24–1.27) | 1.31 (1.23–1.39) | < 0.001 |

Abbreviations: APOE, apolipoprotein E; SD, standard deviation; CI, confidence interval; Aβ, amyloid-β; SUVR, standardized uptake value ratio; –, missing data.

The tau PET subset included 758 participants with a mean (SD) age of 72.3 (6.1) years; 60.2% were women and educational attainment was similar to that of the full sample. APOE genotype frequencies were comparable, with ε33 (48.8%) and ε34 (36.5%) representing the largest groups, followed by ε23 (8.2%), ε44 (3.7%), ε24 (2.6%), and ε22 (0.1%).

In both samples, age differed statistically across APOE genotype groups (amyloid PET sample: p < .001; tau PET subset: p = .002), although the absolute differences were small and unlikely to be clinically meaningful. Sex distribution and education did not differ significantly across groups. Cognitive domain scores were also similar across APOE genotype groups in both datasets (all p ≥ 0.05).

### APOE genotype is associated with amyloid burden and positivity

Descriptive analyses showed clear differences in amyloid burden and amyloid positivity across APOE genotype groups (Table 1; Figure 1A). Mean amyloid levels ranged from 9.73 Centiloids in the ε33 group to 53.20 in the ε44 group, with progressively greater levels observed across ε4-containing genotypes (p < .001) (Table 1; Figure 1A). A similar pattern was observed for amyloid positivity, which ranged from 19.0% in the ε33 group to 72.9% in the ε44 group (p < .001) (Table 1). Pairwise comparisons confirmed higher amyloid levels in ε24, ε34, and ε44 genotype groups compared with ε23 and ε33 groups (all adjusted p < .001) (Figure 1A). Amyloid levels were similar between ε23 and ε33 groups and between ε24 and ε34 groups. Results involving the ε22 group should be interpreted cautiously because of the small sample size.

**Figure 1.**
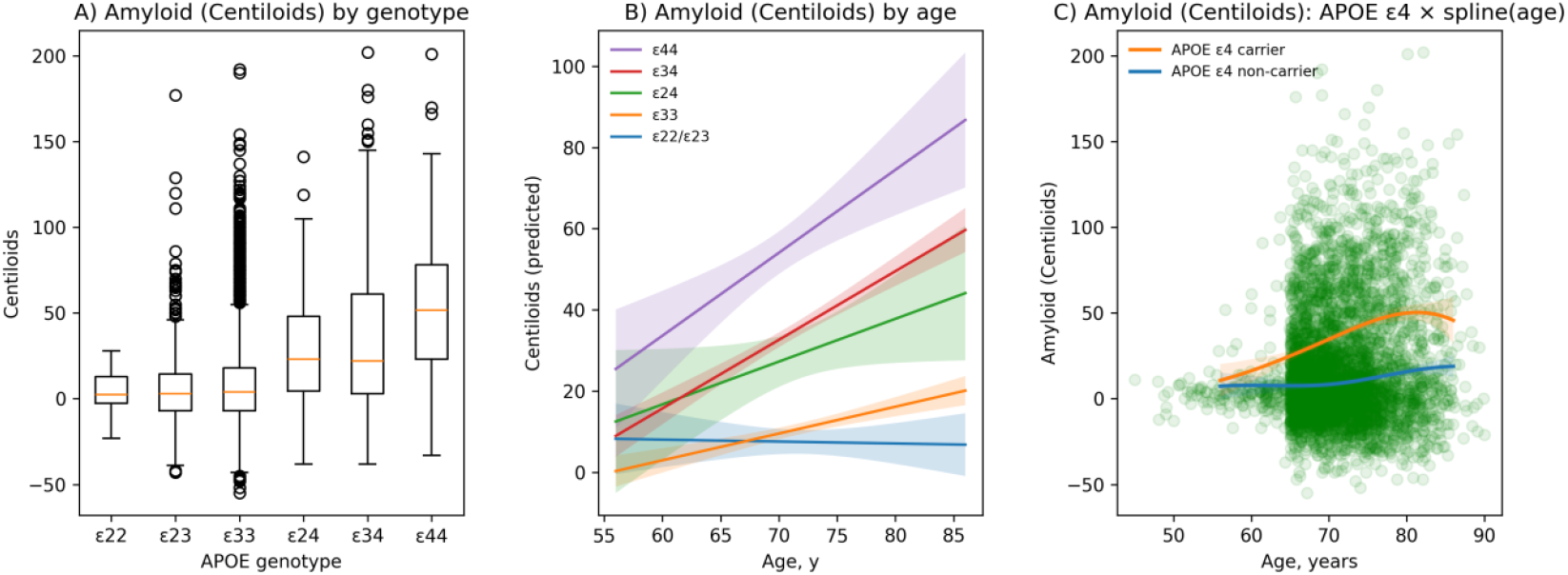
Association between APOE genotype, age, and amyloid burden. (A) Distribution of amyloid burden across APOE genotypes, showing progressively greater amyloid burden across ε4-containing genotypes. (B) Predicted amyloid burden by age according to APOE genotype from covariate-adjusted regression models, demonstrating larger cross-sectional age-associated differences in amyloid burden among ε4-containing genotypes. (C) Spline-based sensitivity analysis showing nonlinear cross-sectional age-associated differences in amyloid burden according to APOE ε4 carrier status.

After adjustment for study cohort, age, sex, and education, APOE genotype remained strongly associated with continuous amyloid burden (LRT χ^2^(4) = 666.40; p < .001). Compared with the ε33 group, ε24 (β = 18.34, p < .001), ε34 (β = 23.850, p < .001), and ε44 (β = 44.82, p < .001) showed higher amyloid burden, while ε22/ε23 showed slightly lower levels (β = −3.46, p = .032). APOE genotype also remained strongly associated with amyloid positivity after covariate adjustment. Relative to ε33, ε24 (OR = 4.18, p < .001), ε34 (OR = 4.54, p < .001), and ε44 (OR = 14.15, p < .001) showed higher odds of amyloid positivity. Amyloid positivity in ε22/ε23 did not differ significantly from ε33 (OR = 0.84, p = .209).

### Age-related differences in continuous amyloid burden by APOE genotype

A significant APOE genotype × age interaction was observed for amyloid burden (ΔAIC = −37.96; LRT χ^2^(4) = 45.96; p < .001), indicating that age-associated differences in amyloid burden varied across APOE genotype groups (Figure 1B). In ε33 group, older age was associated with higher amyloid burden, corresponding to 0.66 Centiloids per year of age. ε22/ε23 groups showed minimal age-associated differences in amyloid burden (−0.05 Centiloids per year). Larger age-associated differences were observed in ε24 (1.05), ε34 (1.69), and ε44 group (2.04), consistent with greater amyloid burden at older ages among ε4-containing genotypes (Figure 1B).

Sensitivity analyses using spline-based age models did not support a significant overall nonlinear association between age and amyloid burden (ΔAIC = 3.71; LRT χ^2^(3) = 2.29; p = .514). However, adding APOE ε4 × spline(age) interaction terms improved model fit substantially (ΔAIC = −32.76; LRT χ^2^(4) = 40.76; p < .001), suggesting nonlinear age-associated differences in amyloid burden between ε4 carriers and non-carriers (Figure 1C).

No significant APOE × sex interactions were observed for either continuous amyloid burden ((LRT χ^2^(4) = 2.40; p = .663) or amyloid positivity (LRT χ^2^(4) = 2.69; p = .610).

### Amyloid burden is associated with tau pathology

Sequential regression models examining global tau burden are presented in Table S1. Descriptive analyses of the tau PET sample showed clear differences in amyloid burden, amyloid positivity, and tau burden across APOE genotype groups (Table 2). Mean amyloid burden ranged from 22.46 Centiloids in ε33 group to 60.61 in ε44 group, with progressively higher amyloid levels across ε4-containing genotypes (p < .001) (Table 2).

Tau burden was higher in Aβ-positive individuals than in Aβ-negative individuals (mean tau SUVR: 1.269 vs. 1.214; Mann–Whitney U = 52761.00; p < .001) (Figure 2A). After adjustment for covariates, amyloid positivity remained associated with higher tau burden (β = 0.0598; p < .001) (Figure 2B). No significant AmyloidPos × age interaction was observed (ΔAIC = 2.00; LRT χ^2^(1) = 0.00; p = .980).

**Figure 2.**
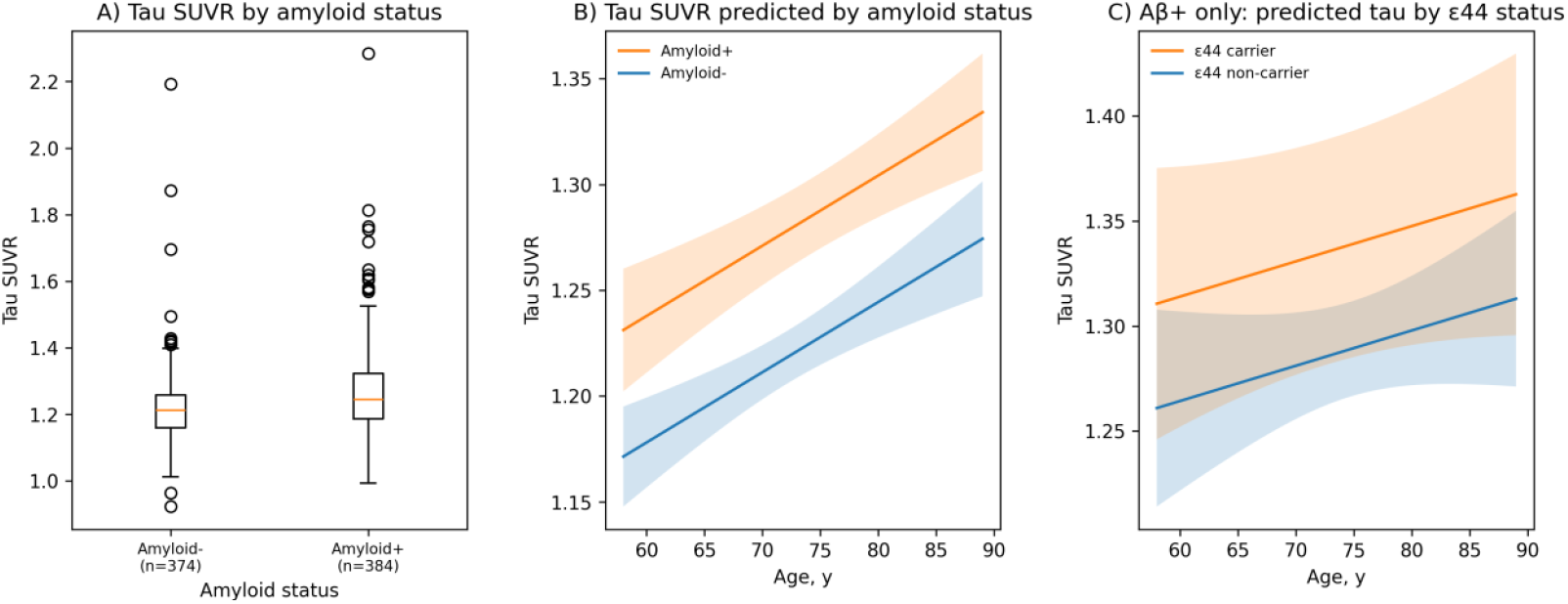
Association between amyloid status, age, APOE ε44 status, and global tau burden. (A) Distribution of global tau SUVR according to amyloid status, showing higher tau burden among amyloid-positive individuals (p < .001). (B) Predicted global tau SUVR by age according to amyloid status from covariate-adjusted regression models, showing higher tau burden across age among amyloid-positive individuals. (C) Among amyloid-positive individuals, predicted global tau SUVR by age stratified by APOE ε44 carrier status, showing modestly higher differences in tau burden after adjustment for continuous amyloid burden and covariates

When amyloid burden was analyzed continuously, higher Centiloid values were associated with greater tau burden after covariate adjustment (β = 0.0012; p < .001). A significant Centiloids × age interaction was observed (ΔAIC = −2.58; LRT χ^2^(1) = 4.58; p = .032). However, spline-based age models did not support significant nonlinear age-related differences in tau burden either before adjustment for amyloid burden (ΔAIC = −0.04; LRT χ^2^(3) = 6.04; p = .109) or after adjustment for continuous amyloid burden (ΔAIC = 1.38; LRT χ^2^(3) = 4.62; p = .202).

No significant AmyloidPos × sex interaction (ΔAIC = 1.89; LRT χ^2^(1) = 0.11; p = .745) or Centiloids × sex interaction (ΔAIC = 0.72; LRT χ^2^(1) = 1.28; p = .257) was observed for tau burden.

### APOE genotype shows a modest association with tau burden

Tau burden also differed across APOE genotype groups (p < .001), with higher values generally observed in ε4 carriers. Mean tau SUVR ranged from 1.21 in ε23 group and 1.23 in ε33 group to 1.26 in ε34 group and 1.31 in ε44 group, with ε24 group showing intermediate values (Table 2).

After adjustment for continuous amyloid burden (Centiloids) and covariates, APOE-related differences in global tau burden were more modest. Compared with ε33 group, only ε44 group had higher tau burden (β = 0.0472; p = .03; LRT χ^2^(1) = 4.74; p = .029). Tau burden in ε24 and ε34 group did not differ significantly from ε33 group. No APOE × age interaction was identified, and spline-based age models similarly showed no evidence of nonlinear APOE × age effects on tau burden. APOE ε44 × sex interaction was observed for global tau burden (ΔAIC = −5.42; LRT χ^2^(1) = 7.42; p = .006).

### APOE-related associations with tau burden among amyloid-positive individuals

Analyses were restricted to amyloid-positive participants to further examine APOE-related differences in tau burden among individuals with elevated amyloid pathology.

Amyloid positivity differed across APOE genotype groups in the tau PET subset, increasing from 27.4% in ε23 group and 37.6% in ε33 group to 70.0% in ε24 group, 68.6% in ε34 group, and 82.1% in ε44 group (p < .001) (Table 2).

Within the amyloid-positive subset, mean tau SUVR was lowest in the ε22/ε23 group (1.206 ± 0.101) and highest in the ε44 group (1.326 ± 0.231), with intermediate values in the ε33 (1.260 ± 0.118), ε24 (1.226 ± 0.144), and ε34 groups (1.278 ± 0.127).

After adjustment for continuous amyloid burden (Centiloids) and covariates, overall APOE genotype was not significantly associated with global tau burden (LRT χ^2^(4) = 5.50; p = .240). Compared with ε33 group, ε44 group showed a borderline association with higher tau burden (β = 0.0549; p = .054; ΔAIC = −1.51; LRT χ^2^(1) = 3.51; p = .061), no significant differences were observed for ε24 or ε34 groups. Figure 2C illustrates predicted global tau burden according to APOE ε44 status within amyloid-positive individuals, showing higher estimated tau SUVR values in ε44 carriers than non-carriers after adjustment for continuous amyloid burden and covariates.

APOE ε44 × sex interaction was observed for global tau burden (ΔAIC = −5.24; LRT χ^2^(1) = 7.24; p = .007).

### Regional tau analyses reveal spatially selective associations with amyloid and APOE ε4

Given the relatively modest APOE-related differences in global tau burden, regional analyses were performed to determine whether these relationships varied across brain regions. Results are presented for APOE ε4 carrier status (Table 3), similar patterns were also observed for ε44 homozygous status.

**Table 3.** Associations between APOE ε4 carrier status and regional tau burden in the tau PET subset.

| Region | APOE $\epsilon 4$ $\beta$ (95% CI) | Unadjusted p | FDR-adjusted p |
| --- | --- | --- | --- |
| Entorhinal cortex | 0.026 (0.005 to 0.047) | .016 | .041 |
| Amygdala | 0.040 (0.018 to 0.062) | < .001 | .003 |
| Inferior temporal cortex | 0.005 (−0.016 to 0.026) | .625 | .637 |
| Inferior parietal cortex | 0.004 (−0.013 to 0.022) | .637 | .637 |
| Precuneus | 0.004 (−0.011 to 0.020) | .596 | .637 |

Abbreviations: APOE, apolipoprotein E; FDR = false discovery rate; CI, confidence interval.

Across all participants, higher continuous amyloid burden was associated with higher regional tau burden in the entorhinal cortex (β = 0.002), amygdala (β = 0.002), inferior temporal cortex (β = 0.001), inferior parietal cortex (β = 0.001), and precuneus (β = 0.001); all FDR-adjusted p < .001. The largest effects were observed in medial temporal regions, particularly the entorhinal cortex and amygdala.

Centiloids × age interactions were identified in the entorhinal cortex, amygdala, inferior parietal cortex, and precuneus (all FDR-adjusted p ≤ .009). No interaction was observed in the inferior temporal cortex.

APOE ε4 carrier status was associated significantly with a higher tau burden in medial temporal regions (Table 3). ε4 carriers had higher tau levels in the entorhinal cortex (β = 0.026; FDR-adjusted p = .041) and amygdala (β = 0.040; FDR-adjusted p = .003). Associations in the inferior temporal cortex, inferior parietal cortex, and precuneus were not significant after FDR correction.

Among amyloid-positive individuals, APOE-related regional tau effects remained significant in medial temporal regions (Table S2). After adjustment for continuous amyloid burden (Centiloids) and covariates, APOE ε4 carrier status was associated with a higher tau burden in the entorhinal cortex (β = 0.047; FDR-adjusted p = .017) and amygdala (β = 0.057; FDR-adjusted p = .009). Associations in the inferior temporal cortex, inferior parietal cortex, and precuneus did not remain significant after FDR correction.

Continuous amyloid burden remained associated with higher tau burden across all examined regions among amyloid-positive individuals, with the largest effects again observed in medial temporal regions.

No APOE ε4 × age interactions were identified after FDR correction. No regional APOE ε4 × sex interactions remained significant after correction.

### Tau burden is selectively associated with memory and language performance

Cognitive domain scores were generally similar across APOE genotype groups in this CU sample (Tables 1 and 2), and APOE genotype was not consistently related to cognitive performance after covariate adjustment.

After adjustment for covariates, APOE genotype, and amyloid burden, higher tau SUVR remained associated with worse memory performance in models using both binary amyloid status (β = −0.497; FDR-adjusted p = .004) and continuous Centiloid measures (β = −0.439; FDR-adjusted p = .012). Similar associations were observed among amyloid-positive individuals (β = −0.464; FDR-adjusted p = .022). Higher tau burden was also associated with lower language performance in models adjusted for binary amyloid status (β = −0.579; FDR-adjusted p = .020) and continuous Centiloid burden (β = −0.610; FDR-adjusted p = .020), although associations were not significant among amyloid-positive individuals after FDR correction. Similar findings were observed across sensitivity analyses (Figures S2–S3). Tau burden was not associated with executive function or visuospatial performance after adjustment (Figures S4– S5).

In the amyloid PET sample, amyloid burden showed modest associations with cognition overall. Higher amyloid burden was related to lower memory performance in both binary amyloid status models (β = −0.050; FDR-adjusted p < .001) and continuous Centiloid models (β = −0.0009; FDR-adjusted p < .001). No significant associations were observed between amyloid burden and executive function, language, or visuospatial performance in either binary or continuous amyloid models.

Within the tau PET subset, amyloid burden remained independently related to lower memory performance after adjustment for tau burden and covariates. No independent differences were identified for executive function, language, or visuospatial performance.

No consistent Tau × Sex or Amyloid × Sex interactions were observed across cognitive domains.

## Discussion

We evaluated relationships among APOE genotype, amyloid burden, tau pathology, and cognition in CU older adults using harmonized data from the ADSP-PHC. APOE ε4 showed strong dose-dependent associations with both amyloid burden and amyloid positivity, with the highest levels observed in ε44 carriers and larger age-related differences among ε4 carriers. Amyloid burden emerged as the primary correlate of tau pathology. The main findings showed that APOE-related differences in global tau burden were comparatively modest and appeared to be driven primarily by ε44 carriers. Regional analyses showed the clearest APOE-related tau differences in medial temporal regions, particularly the entorhinal cortex and amygdala driven by ε4 carriers. Higher tau burden was related mainly to lower memory and language performance, while differences in executive and visuospatial domains were not observed.

APOE ε4 showed a strong dose-dependent relationship with amyloid burden and amyloid positivity, consistent with previous PET studies in CU individuals showing higher amyloid burden and earlier amyloid positivity among ε4 carriers. Ghisays and colleagues [11] reported progressively greater amyloid burden across APOE ε4 gene dose groups in CU adults, with the highest amyloid levels observed in ε44 carriers together with earlier and larger age-related differences in amyloid PET burden. Young and colleagues [12] similarly found that APOE ε4 was associated with greater continuous amyloid burden among amyloid-positive CU individuals, while APOE ε2 showed protective associations with amyloid burden. Ossenkoppele and colleagues [13] further demonstrated in a large multicohort PET study that APOE ε4 carriers status was associated with substantially earlier amyloid positivity in CU individuals. The present findings support earlier PET studies and show a clear APOE dose effect on amyloid burden, with larger age-related differences among ε4-containing genotypes.

APOE-related differences in tau were smaller than those seen for amyloid and were largely limited to medial temporal regions. This pattern is generally consistent with previous tau PET studies in CU individuals. Prior studies have shown that amyloid burden is the strongest predictor of both cross-sectional and longitudinal tau accumulation, while APOE ε4 tends to show more modest independent associations with tau after accounting for amyloid burden, with findings most evident in medial temporal regions and among ε44 carriers [10,11,21]. Young and colleagues [12] further showed that among amyloid-positive CU individuals, APOE ε4 was associated with higher regional tau PET burden after accounting for amyloid burden, with the strongest effects observed in medial temporal regions. Ossenkoppele and colleagues [13] additionally found that APOE ε4 carrier status was associated with an earlier age of tau PET positivity in CU individuals. Our findings align most closely with these regionally selective tau effects: amyloid burden remained the strongest correlate of tau overall, while APOE-related tau differences were smaller and largely limited to the entorhinal cortex and amygdala among APOE ε4 carrier, with somewhat stronger regional associations observed among amyloid-positive individuals.

Cognitive performance was generally similar across APOE genotype groups after adjustment for covariates, suggesting that APOE genotype alone had limited relationships with cognition in this CU sample. Higher tau burden was associated with lower memory and language performance independent of amyloid burden, while associations with executive and visuospatial performance were not observed. This pattern is consistent with the known progression of tau pathology in Alzheimer’s disease, where tau initially accumulates in medial temporal regions involved in episodic memory before extending into broader neocortical regions associated with broader cognitive dysfunction [22]. Previous studies in CU individuals have similarly shown that higher medial temporal tau burden is related mainly to worse memory performance. Weigand and colleagues [14] also reported that medial temporal lobe tau pathology interacted with APOE ε4 status to influence memory performance independently of amyloid burden in older adults without dementia. More recent longitudinal studies support these findings. Dubbelman and colleagues [23] showed that higher medial temporal and neocortical tau burden was associated with faster decline in everyday functioning among CU older adults with elevated amyloid. Together, these findings suggest that greater tau may be more closely related to subtle cognitive decline during preclinical Alzheimer’s disease than APOE genotype itself, with APOE-related cognitive differences likely reflecting the stronger effects of APOE on amyloid accumulation and comparatively smaller relationships with tau pathology.

We did not observe consistent APOE × sex interactions across amyloid burden, regional tau burden, or cognitive outcomes, although a modest APOE ε44 × sex interaction was identified for global tau burden. The largely negative findings should be interpreted cautiously, as the study was restricted to cognitively unimpaired individuals with relatively limited tau pathology, and the tau PET subset was modest in size. Prior studies have reported mixed findings regarding sex differences in tau pathology and tau-related cognitive decline [13,24,25]. Given the limited consistency of the present sex interaction findings, these results should be interpreted cautiously.

Several limitations should be noted. First, the study included only CU older adults, so the findings may not generalize to later stages of Alzheimer’s disease or to clinically impaired populations. Second, because the analyses were cross-sectional, it was not possible to determine the temporal sequence of relationships among amyloid burden, tau pathology, and cognition. Longitudinal studies are needed to clarify how these effects change over time. Third, the sample was drawn from harmonized research cohorts and was dominated by A4 and ADNI participants, which may further limit generalizability to community-based populations. PET and cognitive measures were harmonized across cohorts, but residual differences in recruitment procedures, imaging acquisition, tracer use, and cognitive assessment may remain. Finally, although the overall sample size was large, some APOE genotype groups, particularly ε22, ε24, and ε44, included relatively few participants, limiting statistical power for subgroup analyses and reducing the ability to detect more subtle genotype-specific effects.

## Conclusion

Using harmonized ADSP-PHC data from CU older adults, this study suggests that APOE genotype is most strongly associated with amyloid burden, with smaller and regionally selective associations observed for tau pathology and limited direct associations with cognition. These findings support amyloid burden as the primary APOE-linked biomarker abnormality in CU older adults, while highlighting medial temporal tau as a downstream marker more closely related to subtle cognitive differences.

## Data Availability

All data produced are available online at https://adsp.niagads.org/
Access to these data required registration and approval through controlled-access data repositories

## ACKNOWLEDGMENTS

This study was supported in part by grants from the National Institute of Health (R01AG080635; R01AG095017); the Alzheimer’s Association (SG-24-988292 ISAVRAD); and Cure Alzheimer’s Fund.

The ADSP Phenotype Harmonization Consortium (ADSP-PHC) is funded by NIA (U24 AG074855, U01 AG068057 and R01 AG059716). The harmonized cohorts used in this study within the ADSP-PHC included the Anti-Amyloid Treatment in Asymptomatic Alzheimer’s study (A4 Study), Alzheimer’s Disease Neuroimaging Initiative (ADNI), National Alzheimer’s Coordinating Center (NACC), and Wisconsin Registry for Alzheimer’s Prevention (WRAP). The A4 Study is a secondary prevention trial in preclinical Alzheimer’s disease, aiming to slow cognitive decline associated with brain amyloid accumulation in clinically normal older individuals. The A4 Study is funded by a public-private-philanthropic partnership, including funding from the National Institutes of Health-National Institute on Aging, Eli Lilly and Company, Alzheimer’s Association, Accelerating Medicines Partnership, GHR Foundation, an anonymous foundation and additional private donors, with in-kind support from Avid and Cogstate. The companion observational Longitudinal Evaluation of Amyloid Risk and Neurodegeneration (LEARN) Study is funded by the Alzheimer’s Association and GHR Foundation. The A4 and A4-LEARN Studies are led by Dr. Reisa Sperling at Brigham and Women’s Hospital, Harvard Medical School and Dr. Paul Aisen at the Alzheimer’s Therapeutic Research Institute (ATRI), University of Southern California. The A4 and LEARN Studies are coordinated by ATRI at the University of Southern California, and the data are made available through the Laboratory for Neuro Imaging at the University of Southern California. The participants screening for the A4 Study provided permission to share their de-identified data in order to advance the quest to find a successful treatment for Alzheimer’s disease. We would like to acknowledge the dedication of all the participants, the site personnel, and all of the partnership team members who continue to make the A4 and LEARN Studies possible. Data collection and sharing for this project was funded by the Alzheimer’s Disease Neuroimaging Initiative (ADNI) (National Institutes of Health Grant U01 AG024904) and DOD ADNI (Department of Defense award number W81XWH-12-2-0012). ADNI is funded by the National Institute on Aging, the National Institute of Biomedical Imaging and Bioengineering, and through generous contributions from AbbVie, Alzheimer’s Association, Alzheimer’s Drug Discovery Foundation, Araclon Biotech, BioClinica, Inc., Biogen, Bristol-Myers Squibb Company, CereSpir, Inc., Cogstate, Eisai Inc., Elan Pharmaceuticals, Inc., Eli Lilly and Company, EuroImmun, F. Hoffmann-La Roche Ltd and its affiliated company Genentech, Inc., Fujirebio, GE Healthcare, IXICO Ltd., Janssen Alzheimer Immunotherapy Research & Development, LLC., Johnson & Johnson Pharmaceutical Research & Development LLC., Lumosity, Lundbeck, Merck & Co., Inc., Meso Scale Diagnostics, LLC., NeuroRx Research, Neurotrack Technologies, Novartis Pharmaceuticals Corporation, Pfizer Inc., Piramal Imaging, Servier, Takeda Pharmaceutical Company, and Transition Therapeutics. The Canadian Institutes of Health Research is providing funds to support ADNI clinical sites in Canada. Private sector contributions are facilitated by the Foundation for the National Institutes of Health (www.fnih.org). The grantee organization is the Northern California Institute for Research and Education, and the study is coordinated by the Alzheimer’s Therapeutic Research Institute at the University of Southern California. ADNI data are disseminated by the Laboratory for Neuro Imaging at the University of Southern California. The NACC database is funded by NIA/NIH Grant U24 AG072122. SCAN is a multi-institutional project funded as a U24 grant (AG067418) by the National Institute on Aging. Data collected by SCAN and shared by NACC are contributed by multiple NIA-funded Alzheimer’s Disease Research Centers (ADRCs). The Wisconsin Registry for Alzheimer’s Prevention (WRAP) is supported by R01AG027161 and R01AG054047.

## CONSENT STATEMENT

All human subjects provided written informed consent and permission to share their de-identified data.

## Supplementary Material

**Figure S1.**
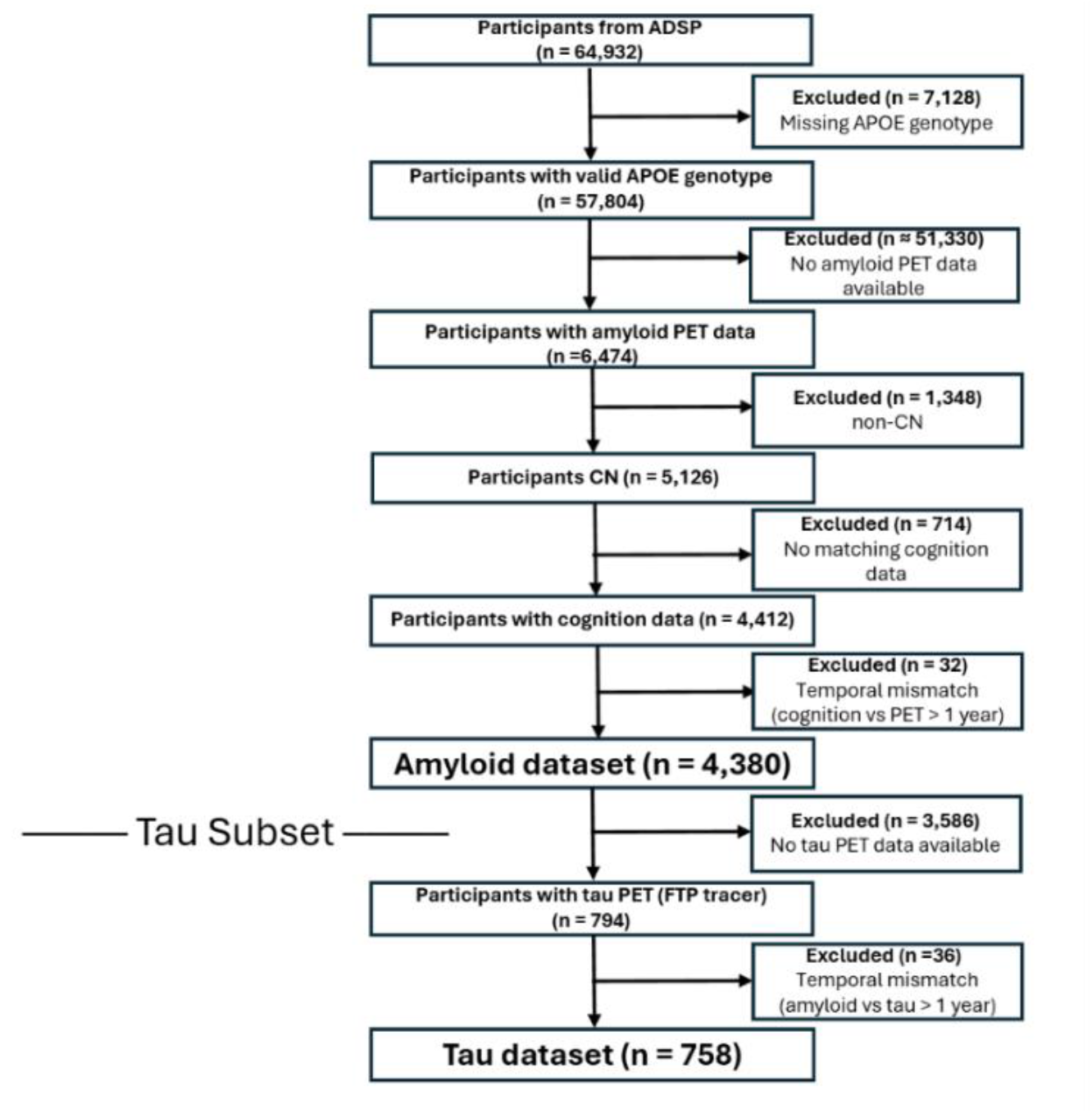
Participant selection flowchart.

**Figure S2.**
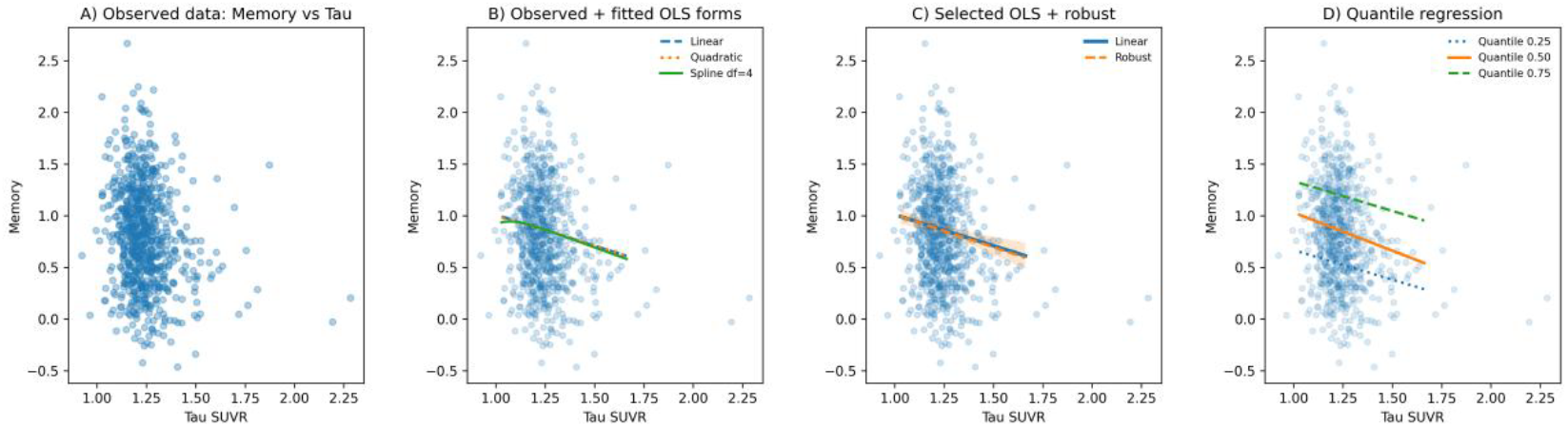
Association between tau SUVR and memory performance across modeling approaches. (A) Observed relationship between tau SUVR and memory scores. (B) Comparison of fitted models, including linear, quadratic, and spline (df = 4), showing no evidence of nonlinearity. (C) Linear and robust regression fits, demonstrating a consistent negative association between tau SUVR and memory. (D) Quantile regression (q = 0.25, 0.50, 0.75), illustrating that the negative association persists across the distribution of memory performance, with similar effects across quantiles.

**Figure S3.**
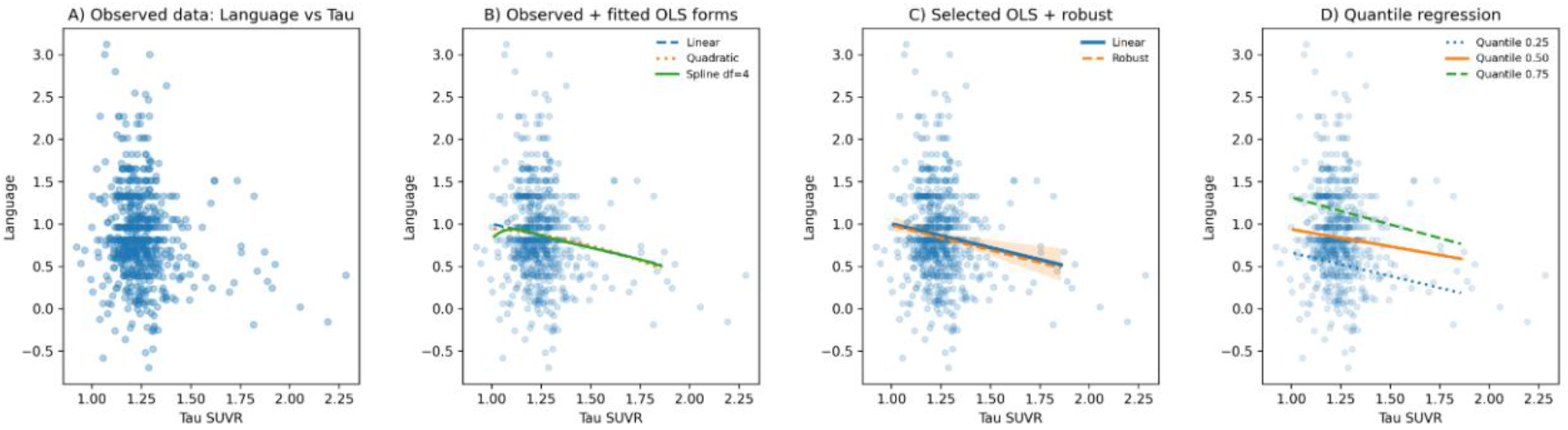
Association between tau SUVR and language performance across modeling approaches. (A) Observed relationship between tau SUVR and language scores. (B) Comparison of fitted models, including linear, quadratic, and spline (df = 4), showing no evidence of meaningful nonlinearity. (C) Linear and robust regression fits, demonstrating a consistent negative association between tau SUVR and language performance. (D) Quantile regression (q = 0.25, 0.50, 0.75), illustrating that the negative association persists across the distribution of language performance, with similar effects across quantiles.

**Figure S4.**
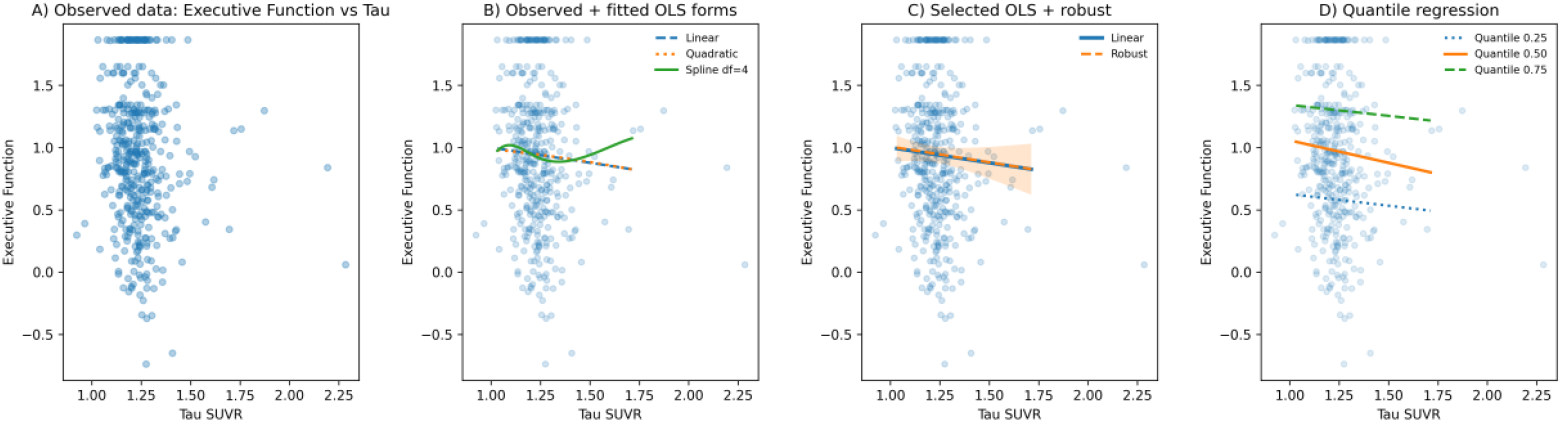
Association between tau SUVR and executive function performance across modeling approaches. (A) Observed relationship between tau SUVR and executive function scores. (B) Comparison of fitted models, including linear, quadratic, and spline (df = 4), showing no evidence of meaningful nonlinearity. (C) Linear and robust regression fits, indicating a weak and inconsistent association between tau SUVR and executive function. (D) Quantile regression (q = 0.25, 0.50, 0.75), demonstrating heterogeneous and generally weak effects across the distribution of executive function performance.

**Figure S5.**
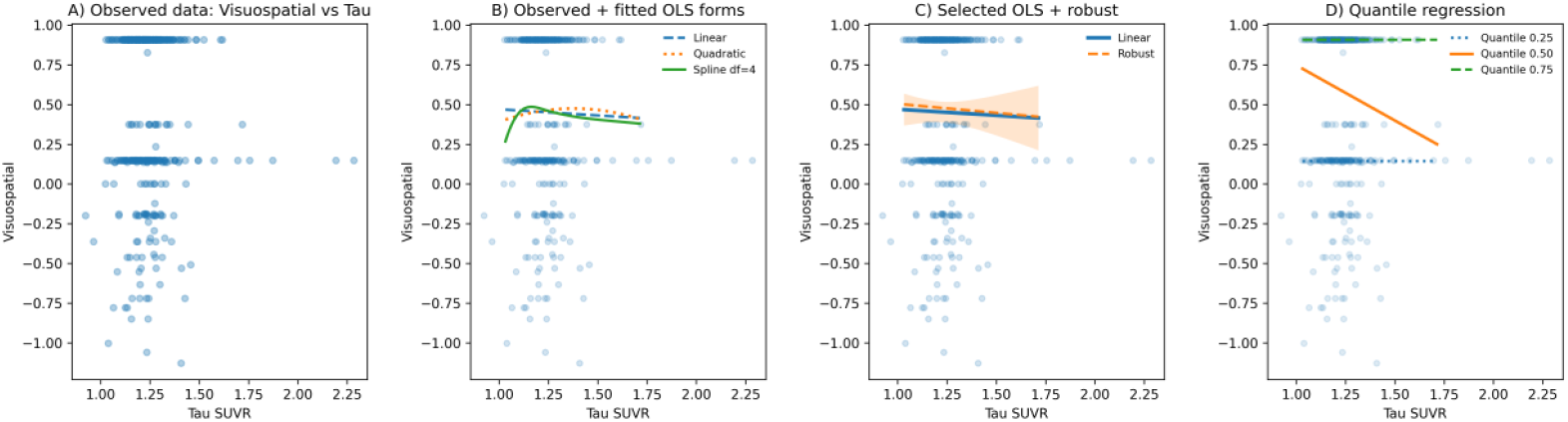
Association between tau SUVR and visuospatial performance across modeling approaches. (A) Observed relationship between tau SUVR and visuospatial scores, showing no clear trend and evidence of outcome discretization. (B) Comparison of fitted models, including linear, quadratic, and spline (df = 4), indicating only minor improvements in fit without a consistent or interpretable association. (C) Spline and robust regression fits, demonstrating the absence of a stable relationship between tau SUVR and visuospatial performance. (D) Quantile regression (q = 0.25, 0.50, 0.75), showing no meaningful association across the distribution of visuospatial performance.

**Table S1.** Regression models evaluating global tau burden.

| Model | Predictor(s) added | $\beta$ (95% CI) | p-value | LRT $\chi^2$(df) | LRT p-value | ΔAIC |
| --- | --- | --- | --- | --- | --- | --- |
| 1 | Base covariates (age, sex, education, cohort) | — | — | — | — | Reference |
| 2a | + Amyloid positivity | 0.060 (0.040 to 0.079) | < .001 | 35.695 (1) | < .001 | -33.695 |
| 2b | + Continuous amyloid burden (Centiloids) | 0.001 (0.001 to 0.001) | < .001 | 102.793 (1) | < .001 | -100.793 |
| 3 | + APOE genotype (overall), adjusted for Centiloids | — | — | 5.227 (4) | .265 | +2.773 |
| 4 | + APOE ε4 carrier status, adjusted for Centiloids | 0.007 (−0.011 to 0.024) | 0.469 | 0.531 (1) | .466 | +1.469 |
| 5 | + APOE ε44 homozygous status, adjusted for Centiloids | 0.047 (0.004 to 0.090) | 0.030 | 4.740 (1) | .029 | −2.740 |

Abbreviations: APOE, apolipoprotein E; CI, confidence interval; LRT, likelihood ratio test; AIC, Akaike information criterion.

**Table S2.**
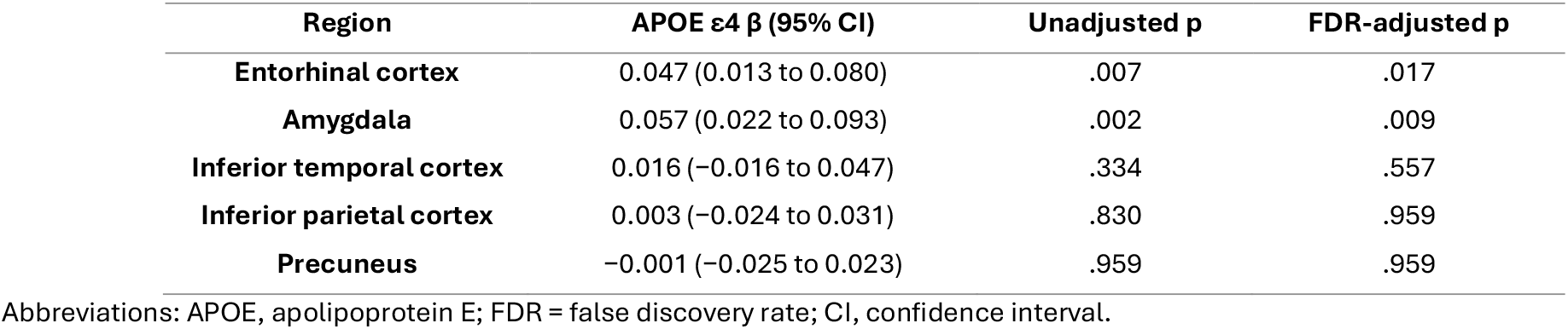
Regional associations between APOE ε4 carrier status and regional tau burden among amyloid-positive individuals.

| Region | APOE ε4 $\beta$ (95% CI) | Unadjusted p | FDR-adjusted p |
| --- | --- | --- | --- |
| Entorhinal cortex | 0.047 (0.013 to 0.080) | .007 | .017 |
| Amygdala | 0.057 (0.022 to 0.093) | .002 | .009 |
| Inferior temporal cortex | 0.016 (−0.016 to 0.047) | .334 | .557 |
| Inferior parietal cortex | 0.003 (−0.024 to 0.031) | .830 | .959 |
| Precuneus | −0.001 (−0.025 to 0.023) | .959 | .959 |

Abbreviations: APOE, apolipoprotein E; FDR = false discovery rate; CI, confidence interval.

